# Bilateral Sequential Theta Burst Stimulation for Multiple-Therapy-Resistant Depression: a naturalistic observation study

**DOI:** 10.1101/2020.02.10.20021782

**Authors:** Amer M. Burhan, James A. Patience, Johannes G.P. Teselink, Nicole M. Marlatt, Sahand Babapoor-Farokhran, Lena Palaniyappan

## Abstract

Depression is a significant health issue with treatment resistance reported in about one third of patients. Treatment resistance results in significant disability, impaired quality of life, and increased healthcare costs. Repetitive transcranial magnetic stimulation (rTMS) is a treatment option for treatment resistant depression (TRD) with an average response rate of around 30%. Theta-burst is a novel rTMS paradigm that has shown promise as a treatment for TRD in some preliminary studies. In a naturalistic design, we evaluated the efficacy and tolerability of bilateral sequential (right then left) prefrontal theta-burst rTMS (bsTBS) in 50 patients with TRD (600 pulses/session, 20 sessions, 100% of resting motor threshold (two patients treated at 80% due to intolerance of 100%), F4/F3 of 10-20-20 EEG localization). Data was collected over 36 months from a specialized academic TMS clinic. Patients had multiple-treatment resistance with at least two failed trials of different antidepressants with 20% also having failed electroconvulsive therapy and 66% having received professional therapy. We found a 28% remission rate (HAMD-17 score of ≤ 7) and a 52% response rate (≥ 50% reduction in HAMD-17) with a 42% reduction in average HAMD-17 score. The treatment was well tolerated, with muscle contractions, mild pain or discomfort, headache, scalp irritation, and changes to vitals being captured as occasional adverse events with two instances of syncope (0.22% of treatments). This naturalistic study shows that bsTBS is a promising paradigm for a multiple-TRD patient population with approximately one-third of treatments achieving remission and over half achieving significant response.

**Previous Publication:** Abstract accepted and study presented at the 2018 Canadian Psychiatric Association Annual Conference, Toronto, Ontario, Canada, September 29, 2018.

## Introduction

Neurostimulation is an increasingly common means of treating depression and is endorsed by the CANMAT guidelines for the treatment of depression (Milev et al., 2016). As depression is associated with asymmetric functional changes (PET and EEG) with hyperactive right and hypoactive left DLPFC activity (Kennedy et al., 1997), rTMS trials often aim to induce either inhibitory plasticity using low frequency pulses to the right or excitatory plasticity using high frequency stimulation to the left DLPFC. Both can also be combined in a bilateral stimulation protocol. Most studies have failed to find evidence of superiority for bilateral vs. unilateral stimulation using standard rTMS (Blumberger et al., 2012; Fitzgerald et al., 2013, 2012; Loo et al., 2003; Pallanti et al., 2010) although bilateral treatment is well tolerated (Berlim et al., 2013; Blumberger et al., 2016). The most widely used, rTMS paradigm for treating depression involves applying 3000 pulses of 10 Hz over 37.5 minutes to the left dorsolateral prefrontal cortex (dlPFC) (George, 2010). These commonly used rTMS paradigms result in remission in 37.1% of patients with treatment resistant depression (TRD; 2.5 ± 2.4 adequate antidepressant trials on average) based on Clinical Global Impression (CGI) scores (Carpenter et al., 2012). While this response rate is encouraging, it is widely recognised that the potential clinical benefits of brain stimulation can be much higher, if the stimulus delivery protocols can be optimised.

Theta burst stimulation (TBS) is a promising patterned form of rTMS, that delivers triplet bursts of energy (50 Hz), at a rate of 5 Hz (Huang et al., 2005). This form of stimulation shares similarities to endogenous neural signaling and can influence neuroplasticity based on how it is administered. When applied continuously (cTBS) it induces long-term depression (LTD) of the target area, inhibiting plasticity; when administered intermittently (iTBS), it induces long-term potentiation (LTP) of the target area, enhancing plasticity (Di Lazzaro et al., 2011; Huang et al., 2005; Suppa et al., 2016). It also has the advantage of reducing administration duration (Chung et al., 2015) from 20-45 minutes to 1-3 minutes. There is preliminary evidence of safety, efficacy, and tolerability of prefrontal theta-burst stimulation in treating depression (Berlim et al., 2017; Cao et al., 2018; Chistyakov et al., 2010; Holzer and Padberg, 2010; Li et al., 2014). Some studies investigated sequential bilateral theta-burst rTMS (bsTBS) in TRD with cTBS to the right dlPFC and then iTBS to the left dlPFC. Some studies were positive (Li et al., 2014; Plewnia et al., 2014) but others were negative (Prasser et al., 2015). The lower number of sessions (15) and low energy level (80% RMT) used might have contributed to these negative results.

Our clinic receives referrals for those with a high level of TRD. Typically, patients have failed several trials of medications and combination therapy. However, the optimal paradigm for these treatments remains a work in progress. In order to optimize the treatment outcome, we chose to target prefrontal areas bilaterally starting with right side cTBS to induce inhibition and pre-condition the left side, followed with left prefrontal area stimulation using iTBS, which is thought to be excitatory. The objective of this study was to assess the response and remission rates of bsTBS in a treatment resistant group of unipolar depressed subjects with ongoing medication and psychotherapy in a naturalistic, retrospective study setting.

## Methods

### Sample and Outcome Measures

This is a retrospective study and was conducted in accordance with the Declaration of Helsinki and was approved by the Western University office of Human Research Ethics Board (WREM) at St. Joseph’s Health Care London. Anonymized data was collected retrospectively, and so informed consent was not required for data acquisition, though all patients consented for the treatment at the outset. Routinely collected measurements of depression and anxiety at baseline and after treatment were used from the on-site hospital-based therapeutic brain stimulation clinic at Parkwood Institute Mental Health Care Building. The assessments used include the 17-item Hamilton Depression Rating Scale (HAMD-17) (Hamilton, 1960), self-report 9-item Patient Health Questionnaire (PHQ-9) (Spitzer et al., 1999), and 7-item Generalized Anxiety Disorder scale (GAD-7) (Spitzer et al., 2006). Data was collected on all adults treated with bsTBS between July 2015 and January 2018. Participants were included if they were between 19 and 80 years old and suffered from treatment resistant unipolar depression as defined by inadequate response (minimal or no improvement) to 2 or more adequate trials of antidepressants from two different drug classes and HAMD-17 ≥ 10.

The primary efficacy measure was defined as a change in HAMD-17 score from baseline to end of treatment. Secondary outcomes included change in PHQ-9 and GAD. A clinical response was defined as ≥ 50% reduction of baseline scores on the HAMD-17 scale and remission was defined by HAMD-17 scores ≤ 7 (Leucht et al., 2013).

### Data analysis

The Statistical Package for the Social Sciences (SPSS 17.0; SPSS, Inc., Chicago IL., USA) was used to run paired t-tests. Scores at baseline for the HAMD-17 (primary outcome), PHQ-9, and GAD were compared to their respective scores at the end of 20 treatment sessions over four weeks. All differences, statistically significant to the p < .05 threshold, were reported. The percentage of the participants that responded (≥ 50% reduction in baseline HAM-D17 score) and remitted (HAMD-17 scores ≤ 7) following treatment were also calculated along with the average percent decrease from baseline to end of treatment for each measure.

### Treatment

In our academic specialized brain stimulation clinic, a TMS Magstim Super Rapid 2 machine (The Magstim Company Ltd™., UK) was used to sequentially apply cTBS at the F4 location of the international 10-20-20 EEG localization system (right dlPFC) followed by iTBS at the F3 location (left dlPFC). Each location received 600 pulses in bursts of 40-50Hz at a rate of 5Hz (theta range) and at 100% of the resting motor threshold (RMT) as established by induction of a visible motor response in the hypothenar hand muscle in 3/5 trials. We allowed the burst frequency to vary between 40-50 Hz to allow treatment at 100% RMT. Tolerability was assessed by any adverse effects during and after each treatment. Twenty sessions were delivered 4-5 days per week. In two cases (4%) the patient could not tolerate the local sensation at 100% energy of the RMT and it was reduced to 80% of RMT.

## Results

Of the subjects included in this naturalistic observational study (n = 50; 47 ± 13.3 years old; 54% female), 10 (20%) had received electroconvulsive therapy and 34 (66%) had received either cognitive behavioural therapy or interpersonal psychotherapy in their lifetime. This was in addition to failing at least two adequate trials of antidepressant medications from different classes in an attempt to treat the current episode, meeting criteria for multiple-TRD (McAllister-Williams et al., 2018).

Within 20 treatment sessions of bsTBS, 14 participants (28%) achieved remission (HAMD-17 scores ≤ 7) and an additional 12 participants responded to treatment (≥ 50% reduction in baseline HAM-D17 score) for a total response rate of 52%. Changes in the average score for each measure from pre-to post-treatment are reported in Figure 1. On average, the HAMD-17 score improved significantly (t(50) = 7.15, p < 0.001) by 43.12% from baseline (M = 22.08, SD = 5.97) to after 20 treatment sessions (M = 12.56, SD = 7.28). The mean PHQ-9 score also improved significantly (t(50) = 6.02, p < 0.001) by 37.05% from 19 ± 4.53 at baseline (M = 19.00, SD = 4.53) to after 20 treatment sessions (M = 11.96, SD = 6.92). Finally, the mean GAD score also improved significantly (t(50) = 7.41, p < 0.001) by 36.71% from baseline (M = 13.62, SD = 4.72) to after 20 treatment sessions (M = 8.62, SD = 6.21). Of the 10 patients who had previously had ECT treatment, seven achieved remission following 20 sessions of treatment. Adverse events were captured for 92% of the sample and are reported in Table 1.

**Table 1:**
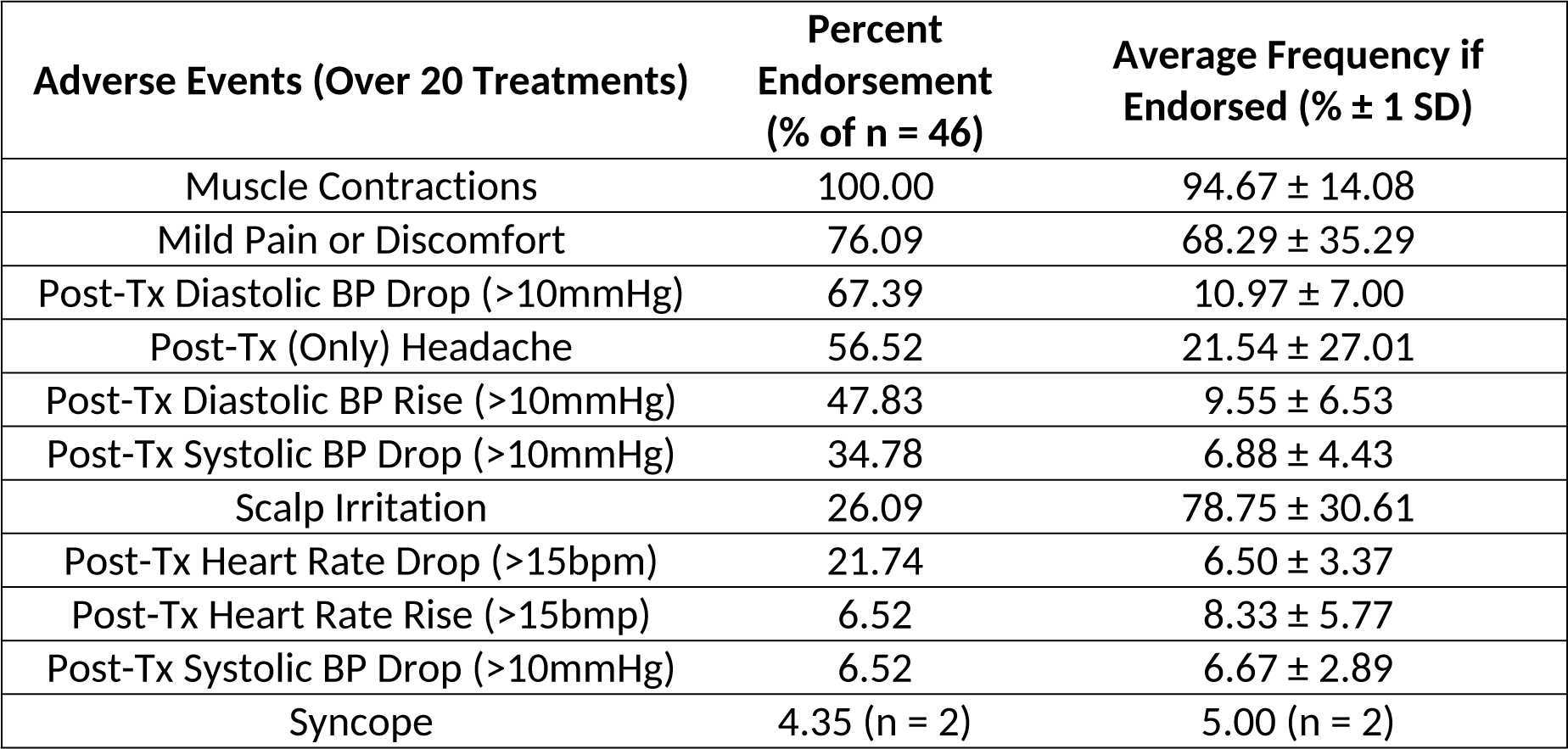
Adverse events experienced in the course of 20 treatments of bilateral sequential theta burst stimulation.

**Figure 1:**
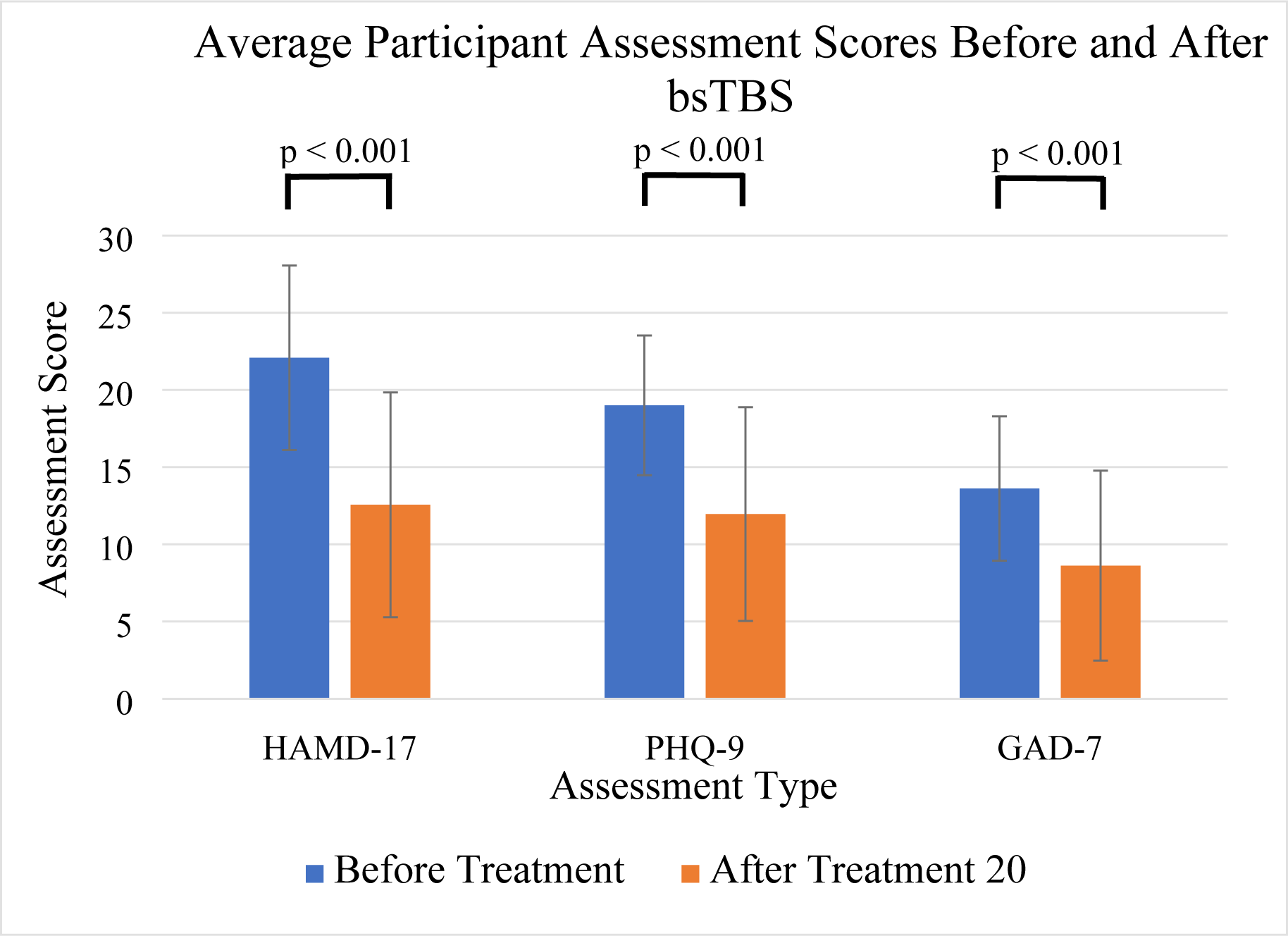
Participant scores on the HAMD-17, PHQ-9, and GAD-7 before and after 20 treatments of bilateral sequential theta burst stimulation.

## Discussion

In this paper we describe the results of a retrospective chart review study completed at an academic therapeutic brain stimulation clinic. All patients included were unipolar major depressive disorder patients with multiple-treatment resistance (at least 2 or more adequate antidepressant trials). Most had failed formal psychotherapy, and some had failed trials of ECT. The paradigm we used in our clinic is novel and involves sequential right then left prefrontal rTMS using continuous then intermittent theta burst stimulation respectively, each 600 pulses. This paradigm was delivered in 10 minutes on average using a Magstim Superrapid 2 machine. The results indicate significant improvement of depressive symptoms on a standardized clinical depression rating scale (HAMD-17) as well as patient rating (PHQ-9) and a standardized assessment of anxiety (GAD-7). Although there have been some studies using bsTBS in this population, to our knowledge this is the first study that does so in a naturalistic tertiary care setting in a highly resistant MDD population.

Our response and remission rates are higher than what was reported in a preliminary meta-analysis of TBS studies (52% vs. 35.6% and 28% vs. 18.6% respectively) (Berlim et al., 2017). Our findings are consistent with the findings in the Li et al. (2014) study in which combined cTBS and iTBS resulted in 66.7% response rate in the group randomized to this arm (n=15). The order of treatment (right pfc cTBS then left iTBS vs. left iTBS then right cTBS) was randomized and the target of stimulation was the junction between Broddmann 9 and 46, which was identified stereotactically based on the individual’s MRI. Each side received 1800 pulses per session. It is likely that providing right then left stimulation using inhibitory (cTBS) then excitatory (iTBS) stimuli, has some advantage over standard unilateral rTMS because of the high efficiency of the theta-burst paradigm in inducing neuroplastic changes and the possibility of affecting a broader distribution of stimulation sites to include right and left frontal-limbic circuits. For instance, TBS has been shown to reliably alter the excitation/inhibition imbalance both at the target and distant cortical sites relevant to the pathophysiology of depression (Iwabuchi et al., 2017; Li et al., 2018). In a previous study comparing 4-weeks of iTBS only with conventional rTMS, there was no notable physiological differences in resting fMRI connectivity or cerebral blood flow after 12 weeks (Iwabuchi et al., 2019). This is likely attributed to a small sample with less severe TRD, resulting in comparable response rates between iTBS and conventional rTMS. The Berlim et al. (2017) meta-analysis also highlights that sequential TBS is the most promising method of TBS treatment for TRD (Berlim et al., 2017). Further mechanistic studies are required to clarify the neural basis of the advantage of bsTBS.

Important limitations to bear in mind, this study was an open-label, retrospective chart review study, and as such is vulnerable to bias. On the other hand, the treatment resistance of the population, rating consistency between clinicians (HAMD-17) and patients (PHQ-9), and the high rate of response and remission, point to a genuine treatment effect. Another limitation is that the Magstim Super Rapid 2 system limits the level of energy delivered using theta-burst at high energy, which led to reducing the burst frequency to between 40-50 Hz. This burst frequency is still however within the gamma burst frequency range and was delivered in a theta-burst triplet pattern at 90-100% of the individual’s resting-motor threshold. Other commercially available machines are now able to deliver the theta-burst frequency at 50 Hz at higher energy levels, which may offer further advantage. Another limitation is related to localization of treatment using an approximation method (Beam F3). While neuro-navigation would have increased the accuracy of targeting, it would not be as easy to use in real-life clinics given the cost and the labour extensive nature of using neuro-navigation systems. In this study we had data on 50 participants who completed 20 sessions of treatment. Other studies have shown that extending the treatment to 30 sessions can add advantage on treatment response (Carpenter et al., 2012). We plan to extend our treatment paradigm to 30 sessions to assess whether this would increase the rate of remission and response. Another factor that needs to be considered is the number of pulses. In Li et al study (Li et al Brain 2014) the number of pulses was higher than our study (1800 vs. 600 pulses right PFC cTBS and left PFC iTBS), which might have accounted to the higher response rate (66.7% compared to 52% respectively). This study did not explore the sustainability of response over time. This is an important issue for future studies.

In summary, for the first time, we report naturalistic data on using bilateral TBS in multi-therapy resistant depressed subjects, demonstrating good tolerability and higher than expected response rates based on unilateral applications. Our data raises the question of the mechanistic differences between unilateral and sequenced bilateral TBS applications. We call for further pragmatic and randomised studies using this approach to demonstrate superior patient benefit.

## Data Availability

The data related to this manuscript are extracted from the clinical charts based on an approved research e5thics board protocol. The data in its anonymous and aggregate form is stored properly at the principle author's site

## Conflict of Interest

Dr. Amer M. Burhan reports personal fees from Janssen Canada for serving on the advisory board for Es-ketamine post trials in TRD and personal fee for providing consultancy to Antheneum medical survey company.

Dr. Lena Palaniyappan reports personal fees from Otsuka Canada, SPMM Course Limited, UK, Canadian Psychiatric Association; book royalties from Oxford University Press; investigator-initiated educational grants from Janssen Canada, Sunovion and Otsuka Canada outside the submitted work. No other authors have conflicts of interest to report.

### Acknowledgments

Joanne Eedy (clinician in the Therapeutic Brain Stimulation Clinic) for supporting the assessment of patients.

Vanessa Eedy (research volunteer) for helping in retrieving charts for the study

Jennifer Speziale and Krista Harloff (Director and manager of the clinic) for supporting chart retrieval cost (in-kind)

## Contributions

*Amer Burhan:* conceptualization, methodology, investigation, resources, writing – original draft, writing - review & editing, project administration

*James Patience:* formal analysis, investigation, data curation, writing – original draft, writing - review & editing, visualization

*Johannes Teselink:* investigation, data curation, writing – review & editing

*Nicole Marlatt:* formal analysis, writing – original draft, writing – review & editing

*Sahand Babapoor-Farokhran:* writing – review & editing

*Lena Palaniyappan:* writing – review & editing, supervision

